# Population Dynamics and Short-Horizon Forecastability of the Burst-Suppression Ratio After Cardiac Arrest: Implications for Closed-Loop Brain-State Monitoring

**DOI:** 10.64898/2026.07.02.26357133

**Authors:** Alon Gorenshtein, Yosef Adiniaev, Tom Liba, Eyal Klang, Oved Daniel

## Abstract

**Objective:** Closed-loop brain-state control uses the burst-suppression ratio as its manipulated variable, assuming the signal is estimable, non-saturated, and predictable over the control horizon. Whether the injured brain’s burst-suppression ratio meets these conditions under routine care is unknown.

**Approach:** We analysed continuous electroencephalography from 607 comatose cardiac-arrest survivors across 5 hospitals in the International Cardiac Arrest Research Consortium database. For each patient we streamed one 24-minute window near 24 hours after return of spontaneous circulation, computed the algorithmic burst-suppression ratio on a 5-second grid, and derived non-saturated occupancy, within-window stability, and short-horizon forecastability. Forecastability was benchmarked against temporal-shuffle surrogates.

**Main results:** The burst-suppression ratio occupied a non-saturated band (0.10 to 0.90) for a median of 0.250 of monitored time (95% CI 0.200 to 0.306) and sat toward suppression (median operating level 0.824). The operating level was stable within the window yet provided little multi-minute predictive information beyond its mean: absolute out-of-sample R^2^ was near zero at 1 to 5 minute horizons, matched temporal-shuffle surrogates, remained near zero in higher-occupancy windows, and was not improved by a sequence model. An autoregressive forecast showed an apparent skill of 0.46 against a persistence baseline, an artifact exposed by the surrogate analysis. Greater occupancy and a lower operating level were associated with good discharge outcome after adjustment (adjusted odds ratio 2.34 and 0.38; both p <= 0.002).

**Significance:** Passive burst-suppression-ratio history provided little multi-minute predictive information beyond the current operating level, although that level was itself estimable and stable within the window. A closed-loop system with a reliable actuator could still regulate the signal by feedback, but model-predictive control from passive dynamics alone would add little.

## Methods

### Study Design and Data Source

This was a retrospective measurement study of continuous electroencephalography (EEG) from comatose survivors of cardiac arrest, using the publicly available International Cardiac Arrest Research Consortium (I-CARE) database version 2.1, distributed through PhysioNet. The database contains baseline clinical information and continuous EEG recordings contributed by seven academic hospitals in the United States and Europe; the public release comprises the 607-patient training partition drawn from five of these sites, with the remaining two sites sequestered in a non-public partition. The study followed the Strengthening the Reporting of Observational Studies in Epidemiology (STROBE) guideline. Primary analyses were prespecified in a design document dated before data extraction; robustness analyses added during revision are identified as such.

The primary objective was descriptive and measurement-focused. We quantified, at fine temporal resolution, the dynamical properties that a closed-loop brain-state controller would assume of the burst-suppression process, observed here without actuation, treating the continuous burst-suppression ratio (BSR) as a candidate control variable. We did not estimate a causal effect and did not develop a prognostic model; the discharge outcome was used only as a secondary construct-validity anchor.

### Cohort

We included all 607 patients in the I-CARE v2.1 public partition. Each patient contributed baseline fields (age, sex, out-of-hospital versus in-hospital arrest, initial shockable versus non-shockable rhythm, targeted temperature management at 33 °C or 36 °C, time from return of spontaneous circulation) and a discharge Cerebral Performance Category (CPC) score. Good neurological outcome was defined as CPC 1 to 2 and poor outcome as CPC 3 to 5, consistent with the database convention. The analysed cohort had a median age of 63 years (IQR, 52-72), 417 of 607 patients (68.7%) were male, 442 (72.8%) had out-of-hospital arrest, 297 (48.9%) had an initial shockable rhythm, and 448 (73.8%) received targeted temperature management at 33 °C. Good outcome occurred in 225 patients (37.1%). The five contributing public sites provided 261, 120, 83, 74, and 69 patients, respectively. A patient was analysable if the selected EEG window yielded a usable BSR series after artifact rejection.

### EEG Selection and Signal Retrieval

EEG was recorded with up to 19 channels in the international 10-20 montage. Native sampling rates ranged from 200 to 2048 Hz across the cohort (most commonly 256, 250, and 500 Hz), and each recording was analysed at its native rate. For each patient we analysed a single window to keep the retrieved data volume tractable: the recording segment whose hour index was closest to 24 hours after return of spontaneous circulation, this timepoint being the most densely covered across the cohort and the canonical post-arrest qEEG assessment window. Ties were resolved toward the later segment. From that segment we retrieved the first 24 minutes of signal using HTTP range requests, decoded the format-16 integer samples to physical microvolts using the per-channel gain and baseline in the WaveForm DataBase header, and deleted the raw signal once the compact feature vector had been written. Retrieved files were verified against the published SHA-256 manifest.

A 24-minute window at a 5-second grid yields 288 burst-suppression-ratio points, of which the first 60% (173 points) train the forecaster and the remainder provide the held-out test region. This gives approximately 104, 92, and 56 forecast origins per window at the 1, 2, and 5 minute horizons in a full-length window. These origins overlap and are not independent, so they do not provide independent evidence within a window; the per-window estimate is least stable at the 5-minute horizon, and all population inference rests on the patient-level bootstrap rather than on within-window origin counts. The single-window design is a deliberate constraint to keep retrieved data volume tractable across the full cohort, and it limits the analysis to a cross-sectional snapshot of within-window dynamics near 24 hours; it does not characterize the 12-to-72-hour evolution of these measures, which we identify as a priority for future work.

### Burst-Suppression Ratio and Preprocessing

Each window was detrended, band-pass filtered between 0.5 and 30 Hz, and notch filtered at the recording-specific utility frequency and its second harmonic. Per 5-second sub-epoch and channel, the signal was rejected as artifact if it saturated (absolute amplitude above 500 µV) or flatlined (standard deviation below 0.1 µV). A channel was scored as suppressed at time points where its sliding root-mean-square amplitude envelope fell below 10 µV for at least 0.5 seconds. A time point was classified as suppressed when a majority of clean channels were suppressed, a spatial-concordance requirement that reduces misclassification of single-channel artifact and of myogenic activity as cerebral suppression. The continuous BSR was computed on a 5-second grid as the fraction of suppressed time within each grid cell, over cells with at least four clean channels. For each window we also recorded the overall clean-channel fraction and the fraction of spectral power above 30 Hz as an electromyographic-contamination index for the sensitivity analysis.

This algorithmic burst-suppression ratio is a voltage-threshold suppression fraction, not an expert-adjudicated American Clinical Neurophysiology Society (ACNS) background classification. It does not incorporate burst morphology, burst duration, highly epileptiform or identical bursts, localization, reactivity, or the continuous, discontinuous, burst-suppression, and suppressed categories of the American Clinical Neurophysiology Society standardized critical care EEG terminology,[12] and the non-saturated band maps only imperfectly onto those categories. We therefore describe properties of the algorithmic signal rather than of expert-classified background. Because a fixed amplitude rule can misclassify technical attenuation, neuromuscular-blockade-related low-voltage recordings, poor electrode contact, or low-amplitude cerebral activity as suppression, we report a prespecified sensitivity analysis over suppression-amplitude thresholds (8, 10, and 12 µV), clean-channel minimums (3, 4, and 6), and the electromyographic veto in eTable S6, re-streamed on a site-balanced subset.

To assess whether the suppression labelling reflected the recording rather than the specific detector design, we additionally compared the primary detector against an independently constructed second detector on the re-streamed site-balanced subset. The second detector replaced the sliding root-mean-square envelope with a peak-to-peak amplitude criterion and the per-channel majority rule with a median amplitude across clean channels, following the classical quantitative burst-suppression definition,[14] with its amplitude threshold calibrated to the primary detector’s suppressed-cell prevalence so that agreement reflected which cells were labelled rather than a prevalence offset. Concordance was summarized by the cell-level Spearman correlation between the primary burst-suppression ratio and the second detector’s amplitude, the Cohen kappa for the binary suppression-dominant cell label, and the patient-level rank concordance of the operating level, each with patient-clustered bootstrap intervals (eTable S12). This is a convergent-validity check across detector designs and is distinct from criterion validity against an expert ACNS reading, which is not claimed here.

### Dynamical Feature Vector

From each patient’s BSR series we computed a prespecified feature vector describing the dynamical properties relevant to closed-loop monitoring. Non-saturated occupancy was the fraction of grid cells with BSR strictly inside a non-saturated band, reported for three bands (0.05-0.95, 0.10-0.90, and 0.20-0.80), capturing the time the process occupied an intermediate, non-saturated range rather than sitting at the continuous or fully suppressed rail. Whether an actuator could in fact move the state within this range is a hypothesis for future interventional work and is not tested here. Regime dwell times were summarized as the mean and coefficient of variation of burst-dominant (BSR < 0.5) and suppression-dominant (BSR⍰0.5) sojourns, with the regime-switch rate expressed per hour. Temporal predictability is detailed below. Operating-level stability over the analysed window was assessed by the augmented Dickey-Fuller (ADF) test, a variance-ratio statistic, and the range of a five-point rolling mean; a window with an ADF probability below .05 was flagged as stationary. Because the ADF test is unreliable on constant or near-constant series, windows with a burst-suppression-ratio standard deviation below 1e-6 were not tested and were excluded from the stationarity denominator, and we report stationarity as stability over the analysed 24-minute window rather than as stationarity of the underlying process.

### Forecastability Model and Surrogate Null

We quantified short-horizon predictability with two forecasters fit on the first 60% of each window and evaluated on the held-out remainder by conditioning on the actual observed history at each origin and iterating the model forward, rather than producing a single dynamic forecast anchored at the training boundary. The first was an autoregressive model with lag order selected by the Akaike information criterion (capped at 10). The second was a two-state switching model in which a Gaussian hidden Markov model assigned each grid cell to a burst-dominant or suppression-dominant regime and a per-regime first-order autoregressive process generated the forecast. The hidden Markov model was fit independently per patient (diagonal covariance, k-means initialization, up to 50 expectation-maximization iterations); states were labeled by their mean burst-suppression ratio; rail-locked regimes with constant predictor values were handled by a zero-slope fallback predicting the regime mean. The model was non-estimable in 16.5% of analysed windows (rail-constant or shorter than 60 points) and added no skill beyond the autoregressive model at any horizon; its absolute out-of-sample R^2^ against the training-window mean was itself markedly negative (median approximately -0.33 to -0.37 across the 1 to 5 minute horizons; eTable S4), so it did not beat predicting the operating-level mean. The primary predictability estimand was the absolute out-of-sample R^2^ against the training-window mean baseline, reported at the one-step (5-second) and 1, 2, and 5 minute horizons. We also report, as a secondary descriptor, the skill over a last-value persistence baseline; we do not treat that quantity as evidence of forecastability, because on a stationary mean-reverting series the persistence baseline is pathologically weak and the persistence-relative skill approaches 0.5 by construction (eMethods S1.5). To test whether any predictability reflected genuine temporal structure rather than the marginal distribution alone, the entire pipeline was repeated on a temporal-shuffle surrogate of each series, which preserves the marginal distribution but destroys autocorrelation; the surrogate absolute R squared served as the null. We validated the estimator against a synthetic first-order autoregressive process and white noise (eMethods S1.5). The switching forecaster was a measurement instrument for predictability and was not used to predict the clinical outcome.

To test whether a more expressive model that pools information across patients could recover predictability the per-patient autoregressive model missed, we also trained a gated-recurrent-unit (GRU) network (one recurrent layer, 24 hidden units) on overlapping two-minute history windows to forecast the burst-suppression ratio 1, 2, and 5 minutes ahead. Patients were pooled and the network was evaluated with leave-one-site-out cross-validation, so it was always tested on a hospital it had not seen, and it was scored by the same absolute out-of-sample R^2^ against each held-out patient’s training-window mean. To confirm the pipeline could detect forecastability when present, we applied it unchanged to synthetic long-memory autoregressive series (first-order coefficient 0.97), whose multi-minute structure is recoverable by construction (eMethods S1.5).

Two further analyses were added during revision rather than prespecified. First, to test whether the near-zero multi-minute forecastability was an artifact of rail-locked windows, we recomputed the forecast metrics within subgroups defined by non-saturated occupancy (greater than 0.20 and greater than 0.40) and by within-window variance (above the cohort median and in the top quartile). Second, to test whether any single hospital drove the headline occupancy and forecastability estimates, we recomputed the cohort medians under leave-one-site-out resampling, dropping each contributing hospital in turn.

### Statistical Analysis

Population distributions of every dynamical metric were summarized as the median with the interquartile range and reported with 95% confidence intervals obtained by patient-level bootstrap resampling with 2,000 resamples; the patient was the unit of resampling so that autocorrelated grid cells within a window did not contribute pseudo-replication. Three metrics were prespecified as the primary set: non-saturated occupancy at 0.10-0.90, the 2-minute absolute out-of-sample R^2^ against the training-window mean, and the fraction of patients with a stationary operating level.

Prespecified subgroup contrasts compared metric distributions across targeted temperature management (33 °C versus 36 °C), arrest location (out-of-hospital versus in-hospital), and initial rhythm (shockable versus non-shockable). Continuous contrasts were tested with the two-sided Mann-Whitney U test and quantified with Cliff’s delta and its 95% bootstrap confidence interval; categorical associations were quantified with Cramér V. As a secondary construct-validity check, the same metrics were compared across good versus poor discharge outcome (Cerebral Performance Category 1 to 2 versus 3 to 5, the latter including death). This comparison was reported with effect sizes and confidence intervals and was explicitly not framed as a prognostic result. To confirm that the outcome associations were not explained by case mix, we also fit exploratory multivariable logistic models of good outcome adjusting for age, initial rhythm, arrest location, temperature-management group, and site; because non-saturated occupancy and the operating level are correlated, each measure was entered in a separate model rather than jointly, and odds ratios are reported per 1.0 increase in the original measure (which ranges from 0 to 1). Across the family of secondary contrasts, P values were adjusted with the Benjamini-Hochberg false discovery rate procedure at a threshold of 0.05. Between-site distributions of the primary metrics were reported across the five available hospitals. All bootstrap confidence intervals were percentile intervals.

Analyses were performed in Python 3.9 with NumPy, SciPy 1.13, statsmodels 0.14, scikit-learn, pandas, hmmlearn 0.3, and PyTorch 2.8 (for the gated-recurrent-unit forecaster). The statistical significance threshold was P < .05.

### Ethics

The I-CARE database is publicly available in deidentified form under a data-use agreement and was collected with approvals at the contributing sites as described in the database documentation. Secondary analysis of this deidentified public dataset did not constitute human-subjects research requiring additional institutional review board approval. No patient consent was required for this secondary analysis. Analysis code and the deidentified derived feature table are available at https://github.com/Alon-Gorenshtein/study_bsr_controllability; the raw EEG remains available to credentialed users through PhysioNet.

## Introduction

Most patients resuscitated from cardiac arrest are comatose at intensive care unit admission, and hypoxic-ischemic brain injury is the leading cause of death in this population.[1,13] Continuous electroencephalography is a standard part of neurological monitoring after arrest, and the burst-suppression pattern, an alternation between high-amplitude bursts and near-isoelectric suppression, is one of its defining features.[2] The burst-suppression ratio, the fraction of time the recording is suppressed, is widely used as a quantitative marker of the depth of cerebral depression.[3]

Beyond its prognostic use, the burst-suppression ratio is the canonical control variable for closed-loop control of brain state. Control-theoretic work has built real-time estimators of burst-suppression probability and demonstrated automated titration of the pattern to a target level under general anesthesia in animals and in silico.[4,5] These systems treat the brain as a plant to be steered: they assume that the controlled variable is observable from the passive signal, that it operates in a non-saturated range where an actuator can move it in either direction, and that it is predictable enough over the control horizon for a model-based controller to act (Figure 1).[6]

**Figure 1.**
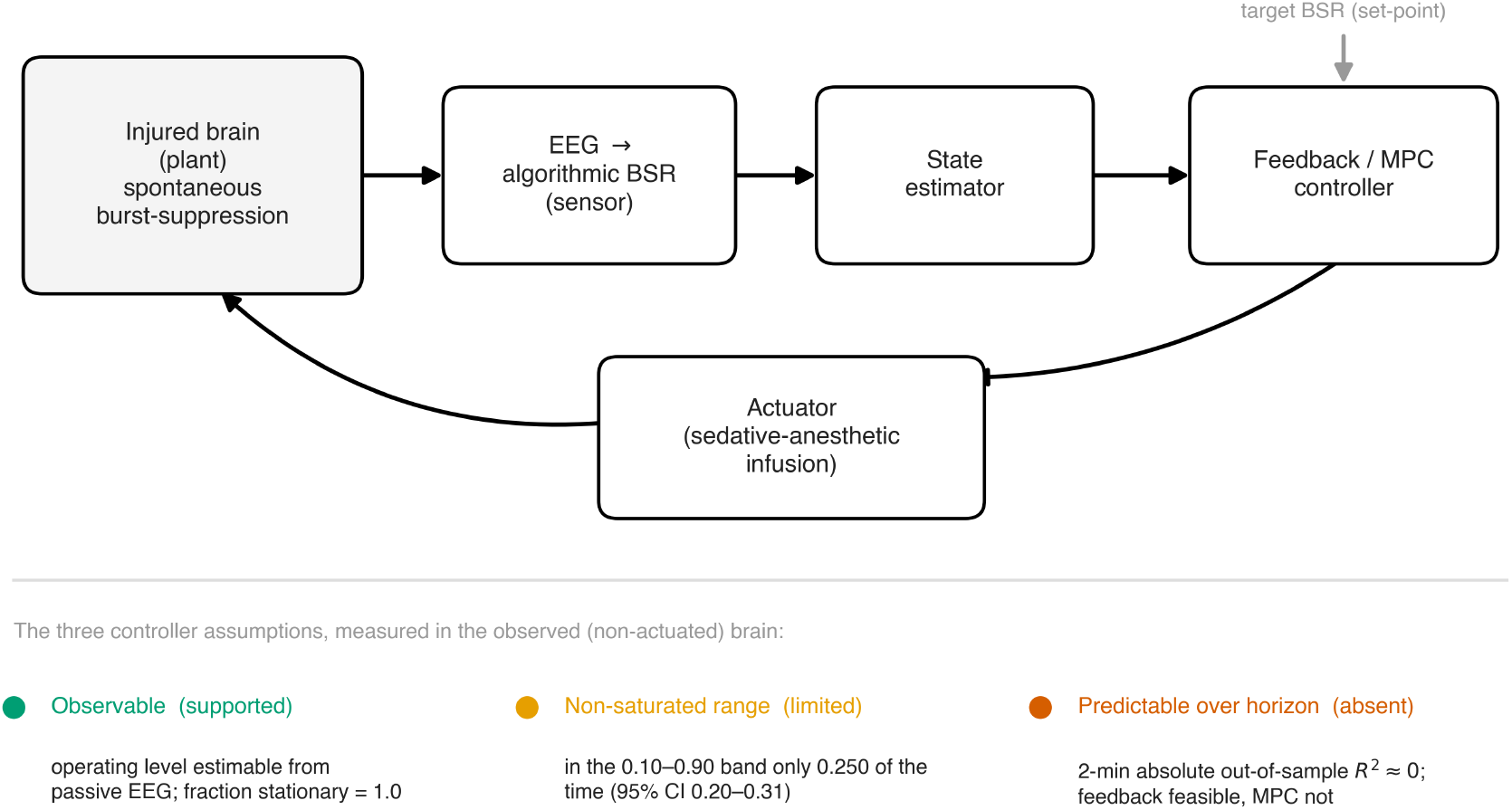
Closed-loop brain-state control loop and the three measured properties of the post-arrest burst-suppression ratio. The burst-suppression ratio is the canonical manipulated variable in closed-loop control of brain state: a sensor estimates it from passive electroencephalography, a state estimator tracks its current value, and a feedback or model-predictive controller drives an actuator (sedative-anesthetic infusion) to hold it at a target, closing the loop through the brain (plant). Every such controller assumes the variable is (i) estimable from the passive signal, (ii) in a non-saturated range where the actuator has headroom, and (iii) predictable over the control horizon. This study measured each assumption in the observed, non-actuated brain: the operating level was estimable and stationary within the window (supported), occupied the non-saturated 0.10-0.90 band for only a median 0.250 of monitored time (limited headroom), and carried near-zero absolute out-of-sample predictive information at 1 to 5 minute horizons (not forecastable). Colors mark the three verdicts. No actuation was performed in this study; the loop is drawn to define the assumptions being tested, not to depict an implemented system.

Whether the observed, non-actuated burst-suppression of the injured human brain actually satisfies these preconditions has not been measured. Prior characterizations of burst-suppression after cardiac arrest have treated it as a static prognostic sign or have modeled coarse multidomain state transitions at 6-hour resolution, without quantifying the fine-resolution dynamical properties a controller would require.[7,8,9] The methods needed to estimate burst and suppression dwell times and transition dynamics exist, but they have not been applied at population scale to ask how often the post-arrest brain presents a non-saturated burst-suppression process and how predictable that process is over a control horizon.[10,11]

In this study, we used continuous electroencephalography from a large multicenter cohort of comatose cardiac-arrest survivors to measure the dynamical properties relevant to closed-loop monitoring of observed, non-actuated burst-suppression. We quantified, at fine temporal resolution, the fraction of time the burst-suppression ratio occupied a non-saturated band, the geometry of its regime sojourns, its within-window stationarity, and its short-horizon forecastability beyond the operating level mean. The study is descriptive and measurement-focused; discharge outcome was used only to test whether the derived measures track a clinically meaningful axis, not to build a prognostic model.

## Results

### Cohort and Signal Quality

The cohort comprised 607 comatose adults resuscitated from cardiac arrest in the I-CARE v2.1 public partition, contributed by 5 academic hospitals (261, 120, 83, 74, and 69 patients at sites A, B, D, E, and F, respectively). The median age was 63 years (IQR 52 to 72), 417 patients (68.7%) were male, 442 (72.8%) had an out-of-hospital arrest, 297 (48.9%) had an initial shockable rhythm, and 448 (73.8%) received targeted temperature management at 33 °C. Good neurological outcome (Cerebral Performance Category 1 to 2) occurred in 225 patients (37.1%).

A single electroencephalography window nearest 24 hours after return of spontaneous circulation was analysed per patient (median window hour 24, IQR 24 to 24). The study flow was as follows: of the 607 patients with a retrievable 24-hour-region segment, 551 (90.8%) met the prespecified signal-quality threshold (clean-channel fraction at least 0.5; median clean fraction 1.00, IQR 0.95 to 1.00) and entered the primary analysis, leaving per-site analysed counts of 252 (site A), 115 (B), 55 (D), 67 (E), and 62 (F). The 56 excluded windows failed the clean-channel threshold. Within the 551 analysed windows, predictability metrics that require a minimum series length and non-zero variance were estimable in 470 to 497 windows (the remainder being too short or rail-constant; see below and eTable S1 for the per-metric denominators). The complete flow is given in eFigure S3.

### The Burst-Suppression Ratio Was Rarely in a Non-Saturated Range

Across the cohort, the burst-suppression ratio spent only a minority of monitored time in a non-saturated range. The median fraction of time with a burst-suppression ratio inside the primary non-saturated band of 0.10 to 0.90 was 0.250 (95% CI 0.200 to 0.306; IQR 0.017 to 0.701) (Figure 2A). Occupancy fell as the band narrowed, to 0.354 (95% CI 0.295 to 0.417) for the 0.05 to 0.95 band and 0.111 (95% CI 0.080 to 0.153) for the 0.20 to 0.80 band. The operating level itself sat toward suppression, with a median per-window mean burst-suppression ratio of 0.824 (95% CI 0.773 to 0.865; IQR 0.440 to 0.985) (Figure 2B). The wide interquartile ranges reflect a bimodal cohort in which many patients were rail-locked at near-continuous or near-suppressed activity while a subset occupied the intermediate band. A small 2048 Hz subgroup (4 analysed windows) was incompatible with the fixed-microvolt detector and returned an occupancy and operating level of exactly 0; these windows were excluded from the descriptive occupancy and operating-level summaries above (see Limitations and eTable S11). Their exclusion left the headline occupancy essentially unchanged (median 0.254 over the remaining 547 windows versus 0.250 over all 551).

**Figure 2.**
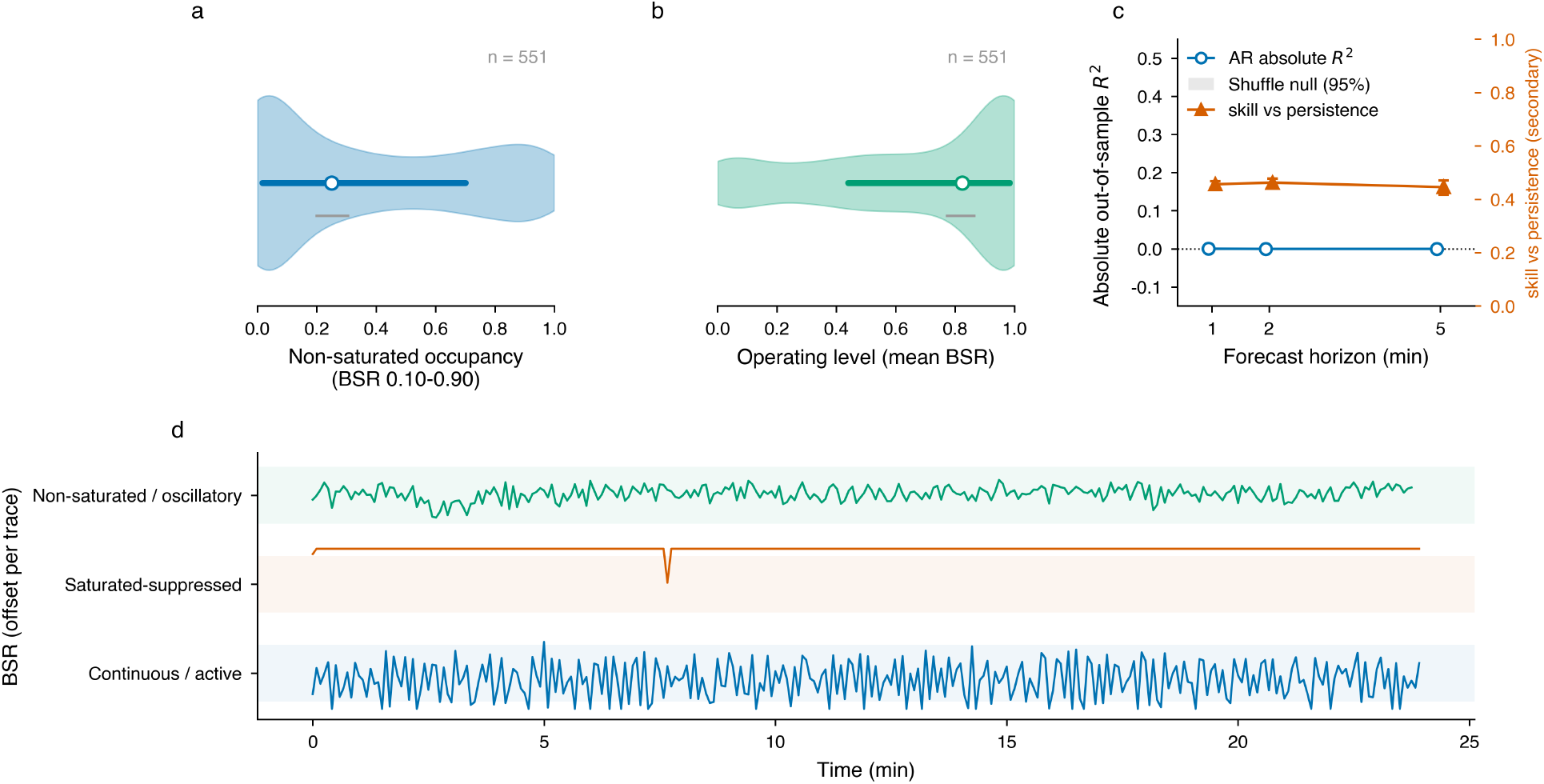
Population distribution and short-horizon forecastability of the algorithmic burst-suppression ratio (n = 551). (a) Distribution of non-saturated occupancy (fraction of the 24-minute window with a burst-suppression ratio inside the 0.10-0.90 band); the cohort spent only a minority of monitored time off the rails. (b) Distribution of the operating level (per-window mean burst-suppression ratio), weighted toward suppression. (c) Short-horizon predictability across 1-, 2-, and 5-minute forecast horizons: the autoregressive absolute out-of-sample R^2^ against the training-window mean (left axis) sits at zero and overlaps the temporal-shuffle null band, while the secondary skill-over-persistence statistic (right axis) sits near 0.46, the persistence-baseline artifact. (d) Three exemplar 24-minute burst-suppression-ratio traces illustrating the continuous/active, saturated-suppressed, and non-saturated/oscillatory regimes.

Regime sojourns were asymmetric. Suppression-dominant episodes had a median dwell time of 90.7 seconds (95% CI 60.3 to 182.5) whereas burst-dominant episodes lasted a median of 5.9 seconds (95% CI 5.0 to 6.7), with regime-switch rates of 10.0 per hour (95% CI 5.0 to 18.2; IQR 0.0 to 142.5).

### The Operating Level Was Stable Within the Window but Carried Little Predictive Information Beyond Its Mean

Within the analysed 24-minute window the burst-suppression operating level was stable. Among the 498 windows with sufficient variance to test (53 windows were not testable, approximately 45 rail-constant plus 8 shorter than the 60-point minimum, and were excluded; 498 + 53 = 551), the augmented Dickey-Fuller test rejected non-stationarity in essentially all (median fraction stationary 1.000, 95% CI 1.000 to 1.000; median p < 0.001), and the variance-ratio statistic was 0.535 (95% CI 0.530 to 0.545; a value near 0.5 indicates white-noise-like increments and a value near 1.0 a random walk, so the series was strongly mean-reverting). We interpret this as stability over the analysed window rather than as stationarity of the underlying 72-hour process, which a single short window cannot establish.

Short-horizon predictability of the burst-suppression ratio beyond its operating-level mean was negligible. The absolute out-of-sample R^2^ of an autoregressive forecast against a training-mean baseline was 0.000 at the 1-minute horizon (95% CI -0.000 to 0.002), -0.000 at 2 minutes (95% CI -0.001 to 0.000), and -0.000 at 5 minutes (95% CI -0.001 to 0.000) (Figure 2C). One-step (5-second) inertia was modest and heterogeneous (median absolute R^2^ 0.013, 95% CI -0.000 to 0.036, IQR -0.056 to 0.204; lag-1 autocorrelation 0.111, 95% CI 0.069 to 0.147). By contrast, the same autoregressive forecast showed an apparent skill over a persistence (last-value) baseline of 0.462 at the 2-minute horizon (95% CI 0.443 to 0.477). This persistence-relative skill was an artifact of the baseline rather than evidence of temporal predictability: on temporally shuffled surrogate series, which retain the marginal distribution but destroy autocorrelation, the absolute out-of-sample R^2^ remained at -0.001 (95% CI -0.001 to 0.000) at 2 minutes, indistinguishable from the real series (Figure 2C). A two-state switching autoregressive model added no skill beyond the autoregressive model; its own absolute out-of-sample R^2^ against the training-window mean was markedly negative at every horizon (median -0.33, -0.37, and -0.36 at 1, 2, and 5 minutes; eTable S4). The near-zero forecastability was not an artifact of applying a Gaussian model to a bounded signal: repeating the analysis on a boundary-adjusted logit transform of the burst-suppression ratio gave a 2-minute absolute R^2^ of -0.000 (IQR -0.008 to 0.011), essentially identical to the untransformed result.

### Forecastability Was Absent in Non-Saturated Subgroups and Under a Pooled Sequence Model

The near-zero multi-minute forecastability was not an artifact of rail-locked patients. Restricting the analysis to the 290 patients who spent more than 20% of monitored time in the non-saturated band (median occupancy 0.68) left the 2-minute absolute out-of-sample R^2^ at 0.000 (95% CI -0.001 to 0.001); restricting further to the 216 patients above 0.40 occupancy (median 0.77) gave 0.001 (95% CI -0.001 to 0.002). Among the 134 patients in the top quartile of within-window variance, short-timescale inertia was clearly present (one-step absolute R^2^ 0.214, 95% CI 0.172 to 0.310) yet 2-minute predictability remained 0.000 (95% CI -0.001 to 0.004), still matching the temporal-shuffle null (eTable S8, eFigure S4). A gated-recurrent-unit network trained across patients with leave-one-site-out validation, which could borrow dynamical structure from the whole cohort, did not beat the operating-level mean either. With each series centered on its own training-window mean before pooling, so the network tested temporal structure rather than the between-patient shift in operating level, its absolute out-of-sample R^2^ sat at essentially zero at every horizon (-0.001, 0.006, and -0.002 at 1, 2, and 5 minutes; eTable S10). Applied to a synthetic long-memory positive control given heterogeneous per-series means that posed the same domain-shift challenge, the identical pipeline recovered clearly positive R^2^ at the horizons it was built to detect (0.42 and 0.13 at 1 and 2 minutes, falling toward zero by 5 minutes as the finite window limits the recoverable signal), confirming it could detect forecastability when present. We report this as suggestive corroboration of the autoregressive null rather than as a capacity-exhausting proof.

### Construct Validity: The Measures Tracked Discharge Outcome

Patients who survived with good discharge outcome spent more time in the non-saturated band and had a lower suppression operating level. Non-saturated occupancy was higher in patients with Cerebral Performance Category 1 to 2 than 3 to 5 (Cliff δ 0.195, 95% CI 0.102 to 0.281; p = 1.2 × 10^-4^, adjusted p = 0.001), and the per-window mean burst-suppression ratio was correspondingly lower (Cliff δ -0.210, 95% CI -0.303 to -0.118; p = 3.4 × 10^-5^, adjusted p = 0.001) (Figure 3). These associations persisted in exploratory multivariable logistic models (each measure entered separately) adjusting for age, initial rhythm, arrest location, temperature-management group, and site: each was independently associated with good outcome (non-saturated occupancy adjusted odds ratio 2.34, 95% CI 1.36 to 4.01, p = 0.002; operating-level adjusted odds ratio 0.38, 95% CI 0.21 to 0.67, p = 0.001; n = 550). No predictability or stationarity metric differed by outcome after correction. These are associations with discharge outcome, reported as construct-validity anchors; discharge Cerebral Performance Category is itself vulnerable to withdrawal-of-life-sustaining-therapy and center-practice variation, so we do not interpret them as prediction of neurological recovery.

**Figure 3.**
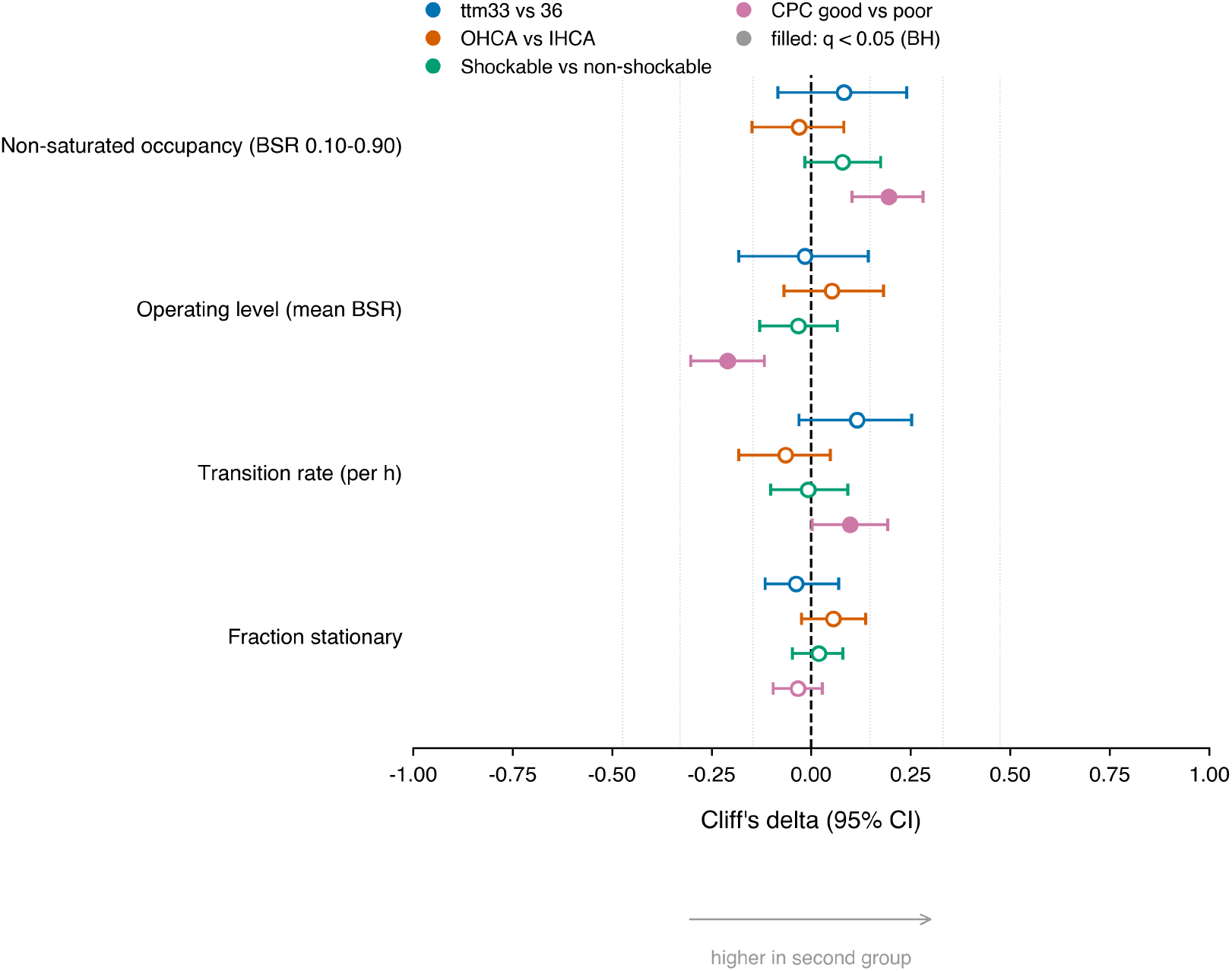
Stratified associations of the dynamical measures. Cliff’s delta with 95% bootstrap confidence intervals for non-saturated occupancy, operating level, transition rate, and fraction stationary across prespecified strata (targeted temperature 33 vs 36 °C, out-of-hospital vs in-hospital arrest, shockable vs non-shockable rhythm) and across good vs poor discharge outcome (a construct-validity anchor, not a prognostic claim). Filled markers indicate associations surviving Benjamini-Hochberg correction (q < 0.05); positive delta indicates a higher value in the first-named group.

### Sensitivity and Cross-Site Analyses

Non-saturated occupancy was similar across the 5 contributing hospitals, with site medians of 0.215 to 0.313 (Figure 4), and the near-zero multi-minute predictability held at every site (median 2-minute absolute R^2^ -0.001 to 0.001). Prespecified subgroup contrasts by targeted temperature management (33 °C versus 36 °C), arrest location (out-of-hospital versus in-hospital), and initial rhythm (shockable versus non-shockable) yielded no difference in any primary metric that survived Benjamini-Hochberg correction across the secondary family. Occupancy estimates were stable across the three non-saturated-band definitions reported above. In leave-one-site-out resampling, the cohort median non-saturated occupancy stayed between 0.249 and 0.253 and the 2-minute absolute out-of-sample R^2^ between -0.001 and 0.000 as each contributing hospital was dropped in turn (eTable S9), so no single site drove either result. In a re-streamed suppression-detector sensitivity analysis spanning the full 24-minute analysis set, median non-saturated occupancy fell monotonically with the suppression-amplitude threshold (0.323, 0.250, and 0.153 at 8, 10, and 12 µV) and was essentially unchanged by the clean-channel minimum, with the rail-weighted pattern preserved at every setting (eTable S6). An independently designed second detector, which replaced the root-mean-square envelope with a peak-to-peak amplitude criterion and the majority rule with a median across clean channels, closely reproduced the labelling on the re-streamed subset (12,354 cells across 45 windows): the cell-level Cohen kappa was 0.87 (95% CI 0.80 to 0.92), the Spearman correlation between the primary burst-suppression ratio and the second detector’s amplitude was 0.86 (95% CI 0.77 to 0.90), cell-level agreement was 94.3%, and patient-level operating levels were concordant (Spearman 0.83; eTable S12); agreement was high at every native sampling rate except 500 Hz, where both detectors placed almost all cells at the suppression rail (99.8% raw agreement), leaving the kappa undefined for want of label variance. All confidence intervals were obtained by patient-level bootstrap resampling with 2000 resamples, which does not account for clustering of patients within hospitals; resampling instead at the hospital-cluster level left the occupancy interval essentially unchanged (median 0.250, 95% CI 0.238 to 0.268 over the 5 sites), consistent with the modest between-site spread seen in the leave-one-site-out analysis.

**Figure 4.**
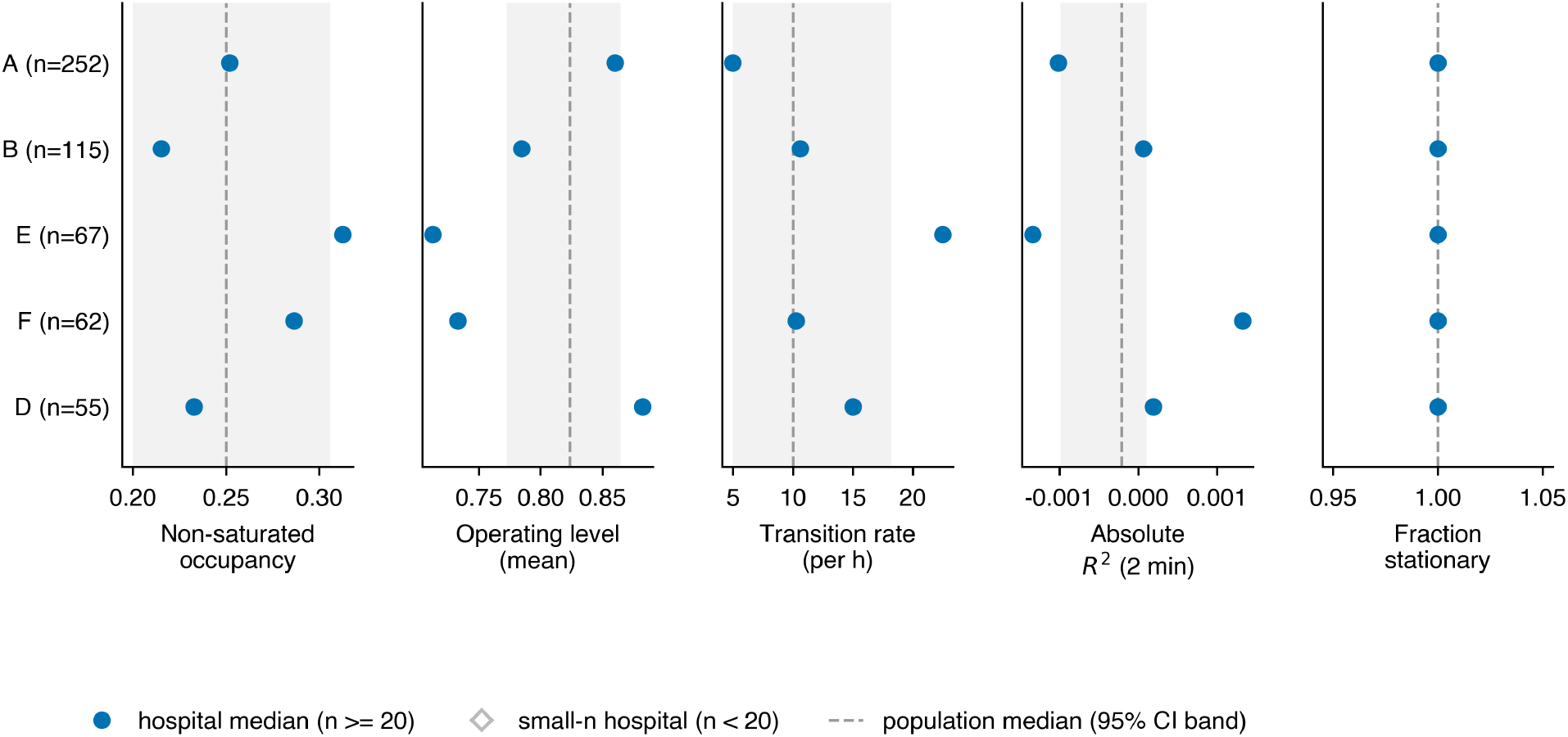
Cross-hospital transportability of the primary metrics. Site medians (one marker per contributing public hospital) for non-saturated occupancy, operating level, transition rate, and fraction stationary, with the population median and its 95% confidence band shaded. Occupancy and the operating level varied modestly across sites; complete within-window stability held at every site.

## Discussion

In a multicenter cohort of comatose cardiac-arrest survivors, the burst-suppression ratio behaved as an operating-level-estimable and within-window-stable signal but not as a forecastable one. For most patients the process sat near a rail, with only about a quarter of monitored time spent in a non-saturated band, and the operating level, although stable within the analysed window, carried almost no information about its own value a few minutes ahead beyond its mean level. State estimation of the current operating level was therefore straightforward, whereas multi-minute prediction had little dynamical structure to exploit.

These properties must be read as properties of the observed signal under routine post-arrest care, not of an isolated injured brain. The I-CARE database does not record sedation, analgesia, neuromuscular-blockade, or detailed temperature exposure, and the burst-suppression ratio at 24 hours reflects the combined influence of anoxic injury, these treatments, antiseizure management, and institutional practice. This confounding is central to interpreting both the operating level distribution and the outcome association, and we accordingly describe non-actuated dynamics under usual care rather than a pharmacologically isolated cerebral state throughout.

This dissociation between stationarity and forecastability is internally coherent. A signal that hovers near a fixed level is easy to estimate and trivially stationary, yet its future deviations from that level can still be close to unpredictable. The negligible absolute out-of-sample R^2^ at the 1 to 5 minute horizons, together with the matching values on temporally shuffled surrogates, indicates that an autoregressive model recovered no held-out temporal information beyond the window mean. The modest and heterogeneous one-step inertia we observed reflects short-timescale persistence of the amplitude envelope rather than exploitable structure at control-relevant horizons. This absence of multi-minute structure held under every check we ran: it persisted in the most variable subgroups, those patients with high non-saturated occupancy or high within-window variance, in whom short-timescale inertia was plainly present; it held under leave-one-site-out resampling; and it was not overcome by a gated-recurrent-unit network that pooled information across the cohort, even though the same network recovered forecastability on synthetic long-memory series. The negative result is therefore unlikely to be an artifact of rail-locked windows, a single site, or a forecaster too simple to capture the dynamics.

These results also expose a measurement pitfall relevant to any predictive-monitoring study in this setting. The same autoregressive forecast that explained no variance beyond the mean nonetheless showed an apparent skill of close to one half over a last-value persistence baseline. For a stationary, mean-reverting series the persistence baseline is pathologically weak, because its error variance approaches twice the signal variance while a model predicting the mean approaches the signal variance, so a skill-over-persistence statistic drifts toward 0.5 by construction. Reporting forecast performance against a persistence baseline alone would have substantially overstated the predictability of the post-arrest electroencephalogram.

Our findings extend prior characterizations of burst-suppression after cardiac arrest. Earlier work has treated the pattern as a static or categorical prognostic sign[3,8] or has modeled multidomain neurophysiology state transitions at 6-hour resolution, reporting that states were largely stationary across individuals.[7] Trajectory analyses of the suppression ratio have clustered patients by their longitudinal course in relation to outcome.[9] None of these quantified continuous-burst-suppression-ratio occupancy of a non-saturated band, regime dwell-time geometry, or short-horizon forecastability framed as control preconditions. The control-theoretic literature that defines burst-suppression probability as the manipulated variable has demonstrated closed-loop titration under anesthesia[4,5] but has not measured whether the observed process in non-titrated patients meets the controller’s assumptions; our results supply that measurement layer and indicate that estimability of the current operating level is the more readily satisfied requirement.

The work has implications for the design of closed-loop neuromodulation in neurocritical care. State estimation of the current operating level is on favorable ground in this population, because the post-arrest burst-suppression ratio is stable over short windows and estimable from passive recording. The absence of multi-minute structure does not imply that a controller would fail, because a feedback controller can regulate a state without long-horizon prediction when the actuator is reliable and the observation loop is fast. The narrower and better-supported conclusion is that model-predictive control based on passive burst-suppression-ratio dynamics alone would have limited advantage over feedback in this population, and that developers should treat predictive horizons here as short and validate any predictive component against distribution-matched null series rather than a persistence baseline. The association between greater non-saturated occupancy, a lower operating level, and good discharge out-come indicates that these passive measures index a clinically meaningful axis, although they describe the observed signal under routine care rather than a pharmacologically isolated cerebral state.

### Study Limitations

This study has several limitations. First, as developed in the interpretation above, the absence of sedation, analgesia, neuromuscular-blockade, and detailed temperature records means the measured burst-suppression ratio cannot be attributed to anoxic injury alone. Second, amplitude-threshold suppression detection cannot fully separate cerebral suppression from neuromuscular-blockade-induced isoelectricity or distinguish genuine bursts from myogenic activity, although we required spatial concordance across channels and quantified high-frequency power to limit this. Third, we analysed a single electroencephalographic window near 24 hours after return of spontaneous circulation per patient to keep retrieved data volume tractable, so the within-patient evolution of these measures over the first 72 hours was not characterized. Fourth, the public I-CARE partition contains 5 of the 7 consortium hospitals, and the cross-site analysis is correspondingly limited. Fifth, the suppression labels were derived from a fixed algorithmic rule rather than expert annotation. Although we report sensitivity across band and threshold definitions and an independently designed second detector closely reproduced the labelling (Cohen kappa 0.87; eTable S12), this establishes convergent validity across detector designs rather than criterion validity, and no expert reference standard was applied; a blinded comparison of the algorithmic non-saturated/rail classification against expert American Clinical Neurophysiology Society background categories is deferred to a future report and is not claimed here. Sixth, the fixed-microvolt root-mean-square suppression detector is sampling-rate dependent: the median operating level rose with the native sampling rate across 200 to 512 Hz (eTable S11), and the 5 recordings sampled at 2048 Hz (4 in the quality-screened set) were incompatible with the fixed-microvolt detector, returning an occupancy and operating level of exactly 0, and were excluded from the descriptive summaries. Because the native sampling rate varies across the contributing hospitals, the cross-site differences in operating level and occupancy partly reflect this native sampling-rate heterogeneity rather than physiology alone. The primary short-horizon forecastability null, however, is sampling-rate invariant, with a 2-minute absolute out-of-sample R^2^ at or near zero in every sampling-rate stratum (eTable S11), so the central conclusion is unaffected. The raw EEG was streamed and deleted once the compact feature vector was written, so resampling all recordings to a common rate was not possible after the fact; a future reextraction at a harmonized rate would let this be tested directly. Seventh, we did not actuate the system, so the sufficiency of any control strategy remains untested; we measured necessary preconditions only.

### Future Directions

Several extensions follow directly. Characterizing the trajectory of non-saturated occupancy and forecastability across the first 72 hours, in the subset of patients with sufficient continuous recording, would establish whether and when the process becomes more or less dynamically variable. Linking these measures to sedation and neuromuscular-blockade records in a cohort where dosing is available would separate the iatrogenic and injury contributions to the observed signal. Replication in the sequestered I-CARE sites and in independent post-arrest cohorts would test cross-site transportability of the fixed suppression-detection rule. Finally, prospective evaluation of a closed-loop operating level-holding controller, with any predictive component benchmarked against shuffled null series, would test whether the favorable estimability we measured translates into control performance at the bedside.

## Conclusion

In comatose survivors of cardiac arrest, the observed, non-actuated burst-suppression ratio was an estimable and stable-within-the-analysed-window but not a multi-minute-forecastable signal, and it occupied a non-saturated band for only a minority of monitored time. A closed-loop controller for this population would face an easy operating-level-estimation problem and a hard predictive-control problem, and predictive monitoring claims in this setting should be validated against distribution-matched null series rather than a persistence baseline.

## Data Availability

All data produced are available online at https://github.com/Alon-Gorenshtein/study_bsr_controllability

## Notes

### Competing Interest Statement

The authors have declared no competing interest.

