## appendix for "Population Dynamics and Short-Horizon Forecastability of the Burst-Suppression Ratio After Cardiac Arrest: Implications for Closed-Loop Brain-State Monitoring"

This Supplementary Information accompanies the main manuscript. It contains the full signal-processing and statistical detail (S1), the complete per-metric and per-stratum results that the main text summarizes (S2), supplementary figures (S3), the analysis codebook (S4), and the code and data availability statement (S5). Primary analyses were prespecified before data extraction; added robustness analyses are identified as such.

Contents

- S1 Supplementary Methods
- S2 Supplementary Tables (S1 to S12)
- S3 Supplementary Figures (S1 to S4)
- S4 Analysis Codebook
- S5 Code and Data Availability

### S1 Supplementary Methods

#### S1.1 Data source and access

The International Cardiac Arrest Research Consortium (I-CARE) database version 2.1 is distributed through PhysioNet under an open data-use agreement. The public partition contains 607 patients drawn from 5 of the 7 contributing hospitals; the remaining 2 sites are held out in a non-public partition. Each patient folder contains a metadata text file and a set of hourly WaveForm DataBase (WFDB) recording segments. Electroencephalography (EEG) was recorded with up to 19 channels in the international 10-20 montage (Fp1, Fp2, F3, F4, C3, C4, P3, P4, O1, O2, F7, F8, T3, T4, T5, T6, Fz, Cz, Pz). Native sampling rates ranged from 200 to 2048 Hz across the cohort (256 Hz in 289 recordings, 250 Hz in 120, 500 Hz in 97, 512 Hz in 56, 200 Hz in 36, 1024 Hz in 4, and 2048 Hz in 5); each recording was analysed at its native rate. Signals were stored as format-16 signed 16-bit integers with a 24-byte data offset and per-channel gain and baseline recorded in the header.

#### S1.2 Window selection and retrieval

For each patient we selected a single recording segment whose hour index relative to return of spontaneous circulation was closest to 24 hours, with ties resolved toward the later segment. This timepoint had the densest coverage across the cohort and corresponds to the conventional post-arrest quantitative-EEG assessment window. We retrieved the first 24 minutes of the selected segment over HTTP range requests rather than downloading whole files, which kept total transferred data near 17 gigabytes for the full cohort. A per-segment wall-clock budget bounded retrieval time so that a single slow network connection could not stall the pipeline. Retrieved files were verified against the published SHA-256 manifest, decoded to physical microvolts, reduced to the compact feature vector described below, and then deleted.

#### S1.3 Burst-suppression ratio computation

Each window was linearly detrended, band-pass filtered between 0.5 and 30 Hz with a fourth-order Butterworth filter applied forward and backward, and notch filtered at the recording-specific utility frequency and its second harmonic. Within each non-overlapping 5-second sub-epoch, a channel was rejected as artifact if its absolute amplitude exceeded 500 microvolts or its standard deviation fell below 0.1 microvolts. The amplitude envelope was computed as a 0.25-second sliding root-mean-square. A channel was scored as suppressed where its envelope fell below 10 microvolts continuously for at least 0.5 seconds. A time point was classified as suppressed when at least half of the clean channels at that time point were suppressed, a spatial-concordance requirement that reduces misclassification of single-channel artifact and of myogenic activity as cerebral suppression. The continuous burst-suppression ratio was computed on a 5-second grid as the fraction of suppressed time within each grid cell, restricted to cells with at least four clean channels. For each window we also recorded the overall clean-channel fraction and the fraction of spectral power above 30 Hz as an electromyographic-contamination index.

#### S1.4 Dynamical feature vector

From each per-patient burst-suppression-ratio series we computed a prespecified feature vector. Non-saturated occupancy was the fraction of grid cells with a value strictly inside a non-saturated band, reported for three band definitions (0.05 to 0.95, 0.10 to 0.90, and 0.20 to 0.80). Regime sojourns were defined by thresholding the ratio at 0.5 into burst-dominant and suppression-dominant runs, summarized by the mean and coefficient of variation of run durations and by the per-hour regime-switch rate. Operating-level stability was assessed with the augmented Dickey-Fuller test, a variance-ratio statistic comparing two-step to one-step increment variance, and the range of a five-point rolling mean. Short-horizon predictability is detailed in S1.5.

#### S1.5 Forecastability estimation and the persistence-baseline artifact

We fit an autoregressive model of order p (selected by the Akaike information criterion, capped at 10) on the first 60 percent of each window. We then produced genuine h-step-ahead forecasts on the held-out remainder by conditioning on the actual observed history at each origin and iterating the autoregressive recursion forward, rather than producing a single dynamic forecast anchored at the training boundary. We report two quantities. Writing y_t for the held-out true value at test index t, f_t for the h-step model forecast, m for the training-window mean, and y_{t-h} for the last observed value at horizon h, the primary quantity is the absolute out-of-sample R squared against the training mean, R2_abs = 1 - sum_t (y_t - f_t)^2 / sum_t (y_t - m)^2, and the secondary quantity is the skill over persistence, skill = 1 - sum_t (y_t - f_t)^2 / sum_t (y_t - y_{t-h})^2.

The two quantities diverge sharply for a stationary, mean-reverting series, and that divergence is the reason the persistence-relative quantity is reported only as a secondary descriptor. For a stationary series the persistence baseline has an error variance that approaches twice the signal variance, while a model that predicts the mean has an error variance that approaches the signal variance, so the skill-over-persistence statistic approaches 0.5 by construction regardless of whether any genuine temporal structure is present. We confirmed this by recomputing both quantities on temporally shuffled surrogate series that preserve the marginal distribution but destroy autocorrelation. On the shuffled surrogates the absolute out-of-sample R squared remained near zero, matching the real series, which establishes that the autoregressive model recovered no held-out predictive information beyond the operating-level mean. We validated the corrected estimator against a synthetic first-order autoregressive process with coefficient 0.6, recovering a one-step absolute R squared of 0.40 against a theoretical value of 0.36, and near zero on white noise. We chose the temporal-shuffle surrogate because, for a linear autoregressive forecaster, a phase-randomized surrogate preserves the autocorrelation the model exploits and therefore cannot test the model; the temporal shuffle destroys it and can. The surrogate analysis is confirmatory rather than decisive: the primary evidence is that the absolute out-of-sample R squared of the real series is itself near zero against the mean baseline. As a further check that the result was not an artifact of applying a Gaussian model to a bounded signal, we repeated the analysis on a boundary-adjusted logit transform of the burst-suppression ratio (clipping to [0.01, 0.99]), obtaining a 2-minute absolute R squared of -0.000 (IQR -0.008 to 0.011), essentially identical to the untransformed result.

As a final test that the near-zero forecastability was not simply an underpowered forecaster, we trained a gated-recurrent-unit network (one recurrent layer, 24 hidden units, Adam optimizer, mean-squared-error loss, batch size 512) on overlapping two-minute history windows pooled across patients, with leave-one-site-out cross-validation so the network was always evaluated on a hospital it had not seen. To isolate temporal structure from the wide between-patient differences in operating level, each series was centered on its own training-window mean before pooling and that mean was supplied as a level feature, so the network predicted deviations from the operating level rather than having to fit a target whose level shifts between patients. It was scored, like the autoregressive model, by the absolute out-of-sample R squared against each held-out patient's training-window mean over that patient's final 40 percent; because centering subtracts a constant from both the target and the baseline, this score is unchanged by the centering and remains the manuscript estimand. On the real cohort the centered network sat at essentially zero at every horizon, confirming no recoverable multi-minute structure beyond the operating level. To confirm the pipeline could detect genuine multi-minute structure under the same domain-shift challenge, we applied it to synthetic first-order autoregressive series with coefficient 0.97 given heterogeneous per-series means spanning the unit interval. The asymptotic per-step ceiling for such a process is the coefficient raised to twice the horizon, but on a finite 288-point window scored on its final 40 percent the achievable ceiling is lower, so we report the recovered numbers rather than quoting that asymptotic value as theory: the network recovered a clearly positive R squared at the 1- and 2-minute horizons (0.42 and 0.13) and fell toward zero by 5 minutes, while on the real cohort it recovered no structure at any horizon (eTable S10, eFigure S4). We frame this analysis as suggestive corroboration of the autoregressive null, not as a capacity-exhausting proof.

##### S1.6 Statistical inference

Population distributions were summarized as the median with the interquartile range. All confidence intervals were obtained by bootstrap resampling of patients with 2000 resamples, with the patient as the resampling unit so that autocorrelated grid cells within a window did not contribute pseudo-replication. Prespecified subgroup contrasts compared metric distributions across targeted temperature management at 33 versus 36 degrees Celsius, out-of-hospital versus in-hospital arrest, and shockable versus non-shockable initial rhythm. Continuous contrasts were tested with the two-sided Mann-Whitney U test and quantified with Cliff delta and its 95 percent bootstrap confidence interval. Across the family of secondary contrasts, raw probabilities were adjusted with the Benjamini-Hochberg false discovery rate procedure. The discharge-outcome contrast (Cerebral Performance Category 1 to 2 versus 3 to 5) was prespecified as a construct-validity anchor. Three metrics were prespecified as the primary set: non-saturated occupancy at 0.10 to 0.90, 2-minute absolute out-of-sample R squared, and the fraction of windows classified stationary. Two robustness analyses were added during revision rather than prespecified: the forecast metrics were recomputed within subgroups defined by non-saturated occupancy and within-window variance (eTable S8, eFigure S4), and the cohort medians of the primary metrics were recomputed under leave-one-site-out resampling, dropping each contributing hospital in turn (eTable S9). Analyses used Python 3.9 with NumPy, SciPy, statsmodels, scikit-learn, pandas, hmmlearn, and PyTorch.

#### S2 Supplementary Tables

Table S1. Full population distribution of every dynamical metric. Values are the cohort median with the bootstrap 95 percent confidence interval and the interquartile range, over windows passing signal-quality screening. The number of windows differs across metrics because some quantities require a minimum series length or non-zero variance.

| Metric | Median | 95% CI | IQR | n |
|---|---|---|---|---|
| Non-saturated occupancy 0.05-0.95 | 0.354 | 0.295, 0.417 | 0.040, 0.796 | 551 |
| Non-saturated occupancy 0.10-0.90 | 0.250 | 0.200, 0.306 | 0.017, 0.701 | 551 |
| Non-saturated occupancy 0.20-0.80 | 0.111 | 0.080, 0.153 | 0.004, 0.504 | 551 |
| Suppression dwell mean (s) | 90.7 | 60.3, 182.5 | 11.0, 1440.0 | 550 |
| Suppression dwell CV | 0.388 | 0.283, 0.474 | 0.000, 0.801 | 515 |
| Burst dwell mean (s) | 5.9 | 5.0, 6.7 | 0.0, 18.1 | 550 |
| Burst dwell CV | 0.488 | 0.428, 0.549 | 0.000, 0.872 | 356 |
| Regime-switch rate (per h) | 10.0 | 5.0, 18.2 | 0.0, 142.5 | 550 |
| Lag-1 autocorrelation | 0.111 | 0.069, 0.147 | -0.011, 0.365 | 499 |
| Absolute R-squared, 1 step (5 s) | 0.013 | -0.000, 0.036 | -0.056, 0.205 | 497 |
| Absolute R-squared, 1 min | 0.000 | -0.000, 0.002 | -0.011, 0.027 | 497 |
| Absolute R-squared, 2 min | -0.000 | -0.001, 0.000 | -0.007, 0.009 | 493 |
| Absolute R-squared, 5 min | -0.000 | -0.001, 0.000 | -0.009, 0.006 | 470 |
| Skill over persistence, 1 step | 0.400 | 0.380, 0.418 | 0.254, 0.492 | 469 |
| Skill over persistence, 2 min | 0.462 | 0.444, 0.477 | 0.332, 0.522 | 466 |
| Augmented Dickey-Fuller p | <0.001 | <0.001 | <0.001 | 498 |

| Metric | Median | 95% CI | IQR | n |
|---|---|---|---|---|
| Variance ratio | 0.535 | 0.530, 0.545 | 0.487, 0.629 | 499 |
| Rolling-mean drift | 0.314 | 0.278, 0.352 | 0.119, 0.547 | 499 |
| Fraction stationary | 1.000 | 1.000, 1.000 | 1.000, 1.000 | 498 |
| Operating level (mean BSR) | 0.824 | 0.773, 0.865 | 0.440, 0.985 | 551 |
| Operating level (median BSR) | 0.893 | 0.837, 0.946 | 0.436, 1.000 | 551 |

Table S2. Prespecified subgroup contrasts. Each cell reports the Cliff delta with its bootstrap 95 percent confidence interval, the raw probability, and the Benjamini-Hochberg adjusted probability across the secondary contrast family. Positive delta indicates a higher value in the first-named group. The discharge-outcome rows are construct-validity anchors, not prognostic claims.

| Contrast | Metric | Cliff δ | 95% CI | Raw p | Adj p | n (a, b) |
|---|---|---|---|---|---|---|
| TTM 33 vs 36 | Occupancy 0.10-0.90 | +0.082 | -0.084, +0.240 | 0.315 | 0.576 | 407, 57 |
| TTM 33 vs 36 | Absolute R-squared 2 min | +0.181 | +0.015, +0.339 | 0.038 | 0.196 | 365, 50 |
| TTM 33 vs 36 | Fraction stationary | -0.038 | -0.116, +0.069 | 0.461 | 0.675 | 370, 50 |
| OHCA vs IHCA | Occupancy 0.10-0.90 | -0.031 | -0.149, +0.082 | 0.619 | 0.743 | 403, 112 |
| OHCA vs IHCA | Lag-1 auto-correlation | -0.128 | -0.253, +0.009 | 0.046 | 0.196 | 364, 104 |
| Shockable vs not | Occupancy 0.10-0.90 | +0.079 | -0.016, +0.175 | 0.120 | 0.361 | 273, 246 |
| Shockable vs not | Absolute R-squared 2 min | +0.091 | -0.013, +0.192 | 0.090 | 0.308 | 248, 217 |
| CPC good vs poor | Occupancy 0.10-0.90 | +0.195 | +0.102, +0.281 | <0.001 | 0.001 | 210, 341 |
| CPC good vs poor | Operating level (mean BSR) | -0.210 | -0.303, -0.118 | <0.001 | 0.001 | 210, 341 |
| CPC good vs poor | Absolute R-squared 2 min | +0.105 | +0.006, +0.212 | 0.049 | 0.196 | 196, 297 |
| CPC good vs poor | Regime-switch rate | +0.098 | +0.002, +0.192 | 0.045 | 0.196 | 210, 340 |

Table S3. Per-hospital distribution of the primary metrics across the 5 contributing public sites. Values are site medians. Occupancy and the operating level varied modestly across sites; the near-zero 2-minute absolute R squared and complete stationarity held at every site.

| Hospital | n | Occupancy 0.10-0.90 | Absolute R-squared 2 min | Fraction stationary | Operating level |
|---|---|---|---|---|---|
| A | 252 | 0.252 | -0.001 | 1.000 | 0.860 |
| B | 115 | 0.215 | 0.000 | 1.000 | 0.785 |
| D | 55 | 0.233 | 0.000 | 1.000 | 0.883 |
| E | 67 | 0.313 | -0.001 | 1.000 | 0.713 |
| F | 62 | 0.287 | 0.001 | 1.000 | 0.733 |

Table S4. Forecastability across horizons with the temporal-shuffle null and the switching-model comparison. The autoregressive absolute out-of-sample R squared is near zero at every horizon and matches the shuffled-surrogate value, while the skill-over-persistence quantity remains near 0.46 as an artifact of the persistence baseline. The two-state switching autoregressive model added no skill beyond the autoregressive model: its own absolute out-of-sample R squared against the training-window mean was markedly negative at every horizon, that is, worse than predicting the operating-level mean.

| Horizon | AR absolute R-squared | 95% CI | Shuffle null R-squared | AR skill over persistence | Switching absolute R-squared (95% CI) | n |
|---|---|---|---|---|---|---|
| 1 min | 0.000 | -0.000, 0.002 | -0.003 | 0.456 | -0.331 (-0.401, -0.259) | 494 |
| 2 min | -0.000 | -0.001, 0.000 | -0.001 | 0.462 | -0.370 (-0.443, -0.306) | 493 |
| 5 min | -0.000 | -0.001, 0.000 | -0.000 | 0.446 | -0.362 (-0.415, -0.282) | 470 |

Table S5. Cohort characteristics. Counts with percentages or medians with the interquartile range, over the 607 patients in the public partition.

| Characteristic | Value |
|---|---|
| Patients | 607 |
| Age (y), median (IQR) | 63 (52-72) |
| Male | 417 (68.7%) |
| Out-of-hospital arrest | 442 (72.8%) |
| Shockable initial rhythm | 297 (48.9%) |
| Targeted temperature 33 °C | 448 (73.8%) |
| Good outcome (CPC 1-2) | 225 (37.1%) |
| Windows passing quality screen | 551 (90.8%) |

| Characteristic | Value |
|---|---|
| Analysed window hour, median (IQR) | 24 (24-24) |

Table S6. Suppression-detector sensitivity across the full analysis set. Median non-saturated occupancy (0.10 to 0.90) and median operating level on the full re-streamed 24-minute analysis set (n = 551 windows passing the clean-channel screen), recomputed under three suppression-amplitude thresholds, three clean-channel minimums, and the primary four-channel rule. The operating level rises and occupancy falls smoothly and monotonically as the amplitude threshold increases, occupancy is essentially unchanged by the clean-channel minimum, and the qualitative finding that the process is rail-weighted with only a minority of time in the non-saturated band is preserved at every setting. This table re-streams the same 24-minute windows used in the primary analysis; the 10 μV row reproduces the primary occupancy estimate as a consistency check.

| Detector setting | Median non-saturated occupancy | Median operating level | n |
|---|---|---|---|
| Suppression threshold 8 μV | 0.323 (IQR 0.022-0.708) | 0.641 | 551 |
| Suppression threshold 10 μV (primary) | 0.250 (IQR 0.017-0.701) | 0.824 | 551 |
| Suppression threshold 12 μV | 0.153 (IQR 0.007-0.635) | 0.918 | 551 |
| Clean-channel minimum 3 | 0.251 (IQR 0.017-0.701) | n/a | 551 |
| Clean-channel minimum 4 (primary) | 0.250 (IQR 0.017-0.701) | n/a | 551 |
| Clean-channel minimum 6 | 0.250 (IQR 0.017-0.701) | n/a | 551 |

Table S7. Exploratory adjusted association with discharge outcome. Multivariable logistic regression of good discharge outcome (Cerebral Performance Category 1 to 2) on each measure, adjusting for age, initial rhythm, arrest location, temperature-management group, and site (n = 550). Odds ratios are per 1.0 increase in the original measure (which ranges from 0 to 1).

| Measure | Adjusted odds ratio | 95% CI | p |
|---|---|---|---|
| Non-saturated occupancy (0.10-0.90) | 2.34 | 1.36, 4.01 | 0.002 |
| Operating level (mean BSR) | 0.38 | 0.21, 0.67 | 0.001 |
| Age (per year) | 0.97 | 0.96, 0.99 | <0.001 |
| Shockable rhythm | 5.41 | 3.46, 8.44 | <0.001 |
| Out-of-hospital arrest | 0.50 | 0.30, 0.84 | 0.009 |

Table S8. Forecastability within non-saturated and high-variance subgroups (added in revision). Forecast metrics recomputed within subgroups defined by non-saturated occupancy and within-window variance, to test whether the near-zero multi-minute predictability was an artifact of rail-locked windows. The absolute out-of-sample R squared is against each window's training mean; the shuffle-null column is the matched temporal-shuffle surrogate at the 2-minute horizon. Short-timescale (one-step) inertia rises with variance, but multi-minute predictability stays at zero and matches the null in every subgroup.

| Subgroup | n | Median occupancy | 1-step | 1 min | 2 min (95% CI) | 5 min | Shuffle null, 2 min |
|---|---|---|---|---|---|---|---|
| All analysable | 493 | 0.306 | 0.015 | 0.000 | -0.000 (-0.001, 0.000) | -0.000 | -0.001 |
| Occupancy > 0.20 | 290 | 0.677 | 0.053 | 0.006 | 0.000 (-0.001, 0.001) | -0.000 | -0.000 |
| Occupancy > 0.40 | 216 | 0.774 | 0.053 | 0.009 | 0.001 (-0.001, 0.002) | 0.000 | -0.001 |
| Variance above median | 268 | 0.686 | 0.094 | 0.006 | 0.000 (-0.001, 0.001) | 0.000 | -0.001 |
| Variance top quartile | 134 | 0.682 | 0.214 | 0.014 | 0.000 (-0.001, 0.004) | -0.000 | -0.001 |
| Occupancy > 0.20 and variance above median | 240 | 0.729 | 0.069 | 0.008 | 0.001 (-0.001, 0.002) | -0.000 | -0.001 |

Table S9. Leave-one-site-out stability of the headline metrics (added in revision). Cohort median non-saturated occupancy and 2-minute absolute out-of-sample R squared recomputed with each contributing hospital dropped in turn. Neither estimate moves materially, so no single site drives either result. The remaining-n column is for occupancy; predictability metrics are estimated on the estimable subset (typically about 90% of the remaining windows).

| Hospital left out | n remaining | Median occupancy (95% CI) | 2-min absolute $R^2$ (95% CI) |
|---|---|---|---|
| None (full cohort) | 551 | 0.250 (0.198, 0.306) | -0.000 (-0.001, 0.000) |
| A | 299 | 0.250 (0.188, 0.333) | 0.000 (-0.000, 0.001) |
| B | 436 | 0.253 (0.191, 0.316) | -0.000 (-0.001, 0.000) |
| D | 496 | 0.252 (0.198, 0.313) | -0.000 (-0.001, 0.000) |
| E | 484 | 0.249 (0.191, 0.302) | -0.000 (-0.001, 0.000) |
| F | 489 | 0.250 (0.191, 0.302) | -0.001 (-0.001, 0.000) |

Table S10. Pooled gated-recurrent-unit forecaster with per-patient centering and a synthetic positive control (added in revision; updated to remove an operating-level domain-shift confound). A GRU network was trained across patients with leave-one-site-out validation and scored by absolute out-of-sample R squared against each held-out patient's training-window mean. Each series was centered on its own training-window mean before pooling, with that mean supplied as a level feature, so the network tests temporal structure rather than the between-patient shift in operating level; subtracting a constant from both the target and the baseline leaves the absolute R squared against the training mean unchanged, so the score remains the manuscript estimand. On the real cohort the centered GRU sits at essentially zero at every horizon, confirming no recoverable multi-minute temporal structure beyond the operating-level mean. Applied to a synthetic long-memory autoregressive control (first-order coefficient 0.97) given heterogeneous per-series means that pose the same domain-shift challenge, the same pipeline recovers clearly positive R squared at the 1- and 2-minute horizons, confirming it detects multi-minute structure when present; the synthetic R squared falls toward zero by 5 minutes because the finite 288-point window scored on its final 40% caps the recoverable signal below the asymptotic ceiling. This table is suggestive corroboration of the autoregressive null, not a capacity-exhausting proof.

| Forecast horizon | Real cohort absolute $R^2$ (95% CI) | n | Synthetic AR(1) $\varphi =$ 0.97, heterogeneous means, absolute $R^2$ (95% CI) |
|---|---|---|---|
| 1 min | -0.001 (-0.005, 0.006) | 493 | 0.424 (0.375, 0.451) |
| 2 min | 0.006 (0.003, 0.009) | 491 | 0.126 (0.061, 0.170) |
| 5 min | -0.002 (-0.004, -0.001) | 469 | -0.055 (-0.095, -0.004) |

Table S11. Sampling-rate-stratified sensitivity of operating level, occupancy, and 2-minute forecastability (added in revision). For each native sampling rate present in the quality-screened cohort (n = 551), the table reports the number of analysed windows, the median non-saturated occupancy (0.10-0.90), the median operating level (mean burst-suppression ratio), and the median 2-minute absolute out-of-sample R squared. The operating level and occupancy vary systematically with the sampling rate across 200 to 512 Hz, reflecting the sampling-rate dependence of the fixed-microvolt root-mean-square suppression detector, and the 5 windows recorded at 2048 Hz (4 in the quality-screened set) show an outright detector breakdown (occupancy and operating level of exactly 0). The primary 2-minute forecastability null, by contrast, holds at every sampling rate: the median absolute R squared is at or near zero in every stratum. The forecast metric is non-estimable in the 2048 Hz subgroup because the detector returns a constant (zero-variance) series.

| Native sampling rate (Hz) | n | Median occupancy 0.10-0.90 | Median operating level | Median absolute $R^2$ 2 min |
|---|---|---|---|---|
| 200 | 26 | 0.201 | 0.508 | -0.001 |
| 250 | 115 | 0.215 | 0.785 | 0.000 |
| 256 | 263 | 0.285 | 0.845 | -0.000 |
| 500 | 96 | 0.132 | 0.914 | -0.002 |
| 512 | 45 | 0.448 | 0.777 | 0.001 |
| 1024 | 2 | 0.177 | 0.816 | -0.003 |
| 2048 (detector breakdown) | 4 | 0.000 | 0.000 | not estimable |

Table S12. Second-detector concordance (convergent validity, added in revision). On the re-streamed site-balanced subset (45 windows, 12,354 quality-screened 5-second cells), the primary sliding root-mean-square detector was compared with an independently designed second detector that used a peak-to-peak amplitude criterion and a median-across-clean-channels rule, with the second detector's amplitude threshold calibrated to the primary detector's suppressed-cell prevalence (0.683) so that agreement reflected which cells were labelled rather than a prevalence offset. This is convergent validity across detector designs, not criterion validity against an expert reading. Confidence intervals are patient-clustered bootstrap intervals. Agreement was high overall and at every native sampling rate except 500 Hz, where both detectors placed almost all cells at the suppression rail, leaving no label variance and an undefined kappa despite 99.8% raw agreement (the kappa paradox under extreme prevalence).

| Concordance measure | Value (95% CI) |
|---|---|
| Cell-level Cohen kappa, suppression-dominant cell | 0.87 (0.80, 0.92) |
| Cell-level Spearman, BSR vs negative peak-to-peak amplitude | 0.86 (0.77, 0.90) |
| Cell-level raw agreement | 94.3% |
| Patient-level operating-level Spearman | 0.83 |

| Native sampling rate (Hz) | Cells | Cohen kappa | Raw agreement |
|---|---|---|---|
| 200 | 2302 | 0.97 | 98.7% |
| 250 | 2280 | 0.78 | 89.2% |
| 256 | 4660 | 0.82 | 94.1% |
| 500 | 1108 | undefined (near-saturation) | 99.8% |
| 512 | 2004 | 0.84 | 92.6% |

#### S3 Supplementary Figures

Figure S1. Distribution of the per-patient operating level and non-saturated occupancy. The operating level is heavily weighted toward suppression and occupancy is bimodal, with many patients rail-locked and a subset in the intermediate band. (Rendered from the population digest; see main Figure 2A and 2B.)

Figure S2. Forecastability against the temporal-shuffle null at the 1, 2, and 5 minute horizons. The absolute out-of-sample R squared overlaps the shuffled-surrogate band at zero, while the skill-over-persistence quantity on the secondary axis sits near 0.46. (See main Figure 2C.)

Figure S3. Study flow. Of 607 patients in the public partition with a retrievable 24-hour-region segment, 551 (90.8%) passed the clean-channel signal-quality threshold and entered the primary analysis (per-site analysed counts: A 252, B 115, D 55, E 67, F 62); 56 windows were excluded for failing the threshold. Within the 551 analysed windows, predictability metrics were estimable in 470 to 497 windows, the remainder being too short or rail-constant.

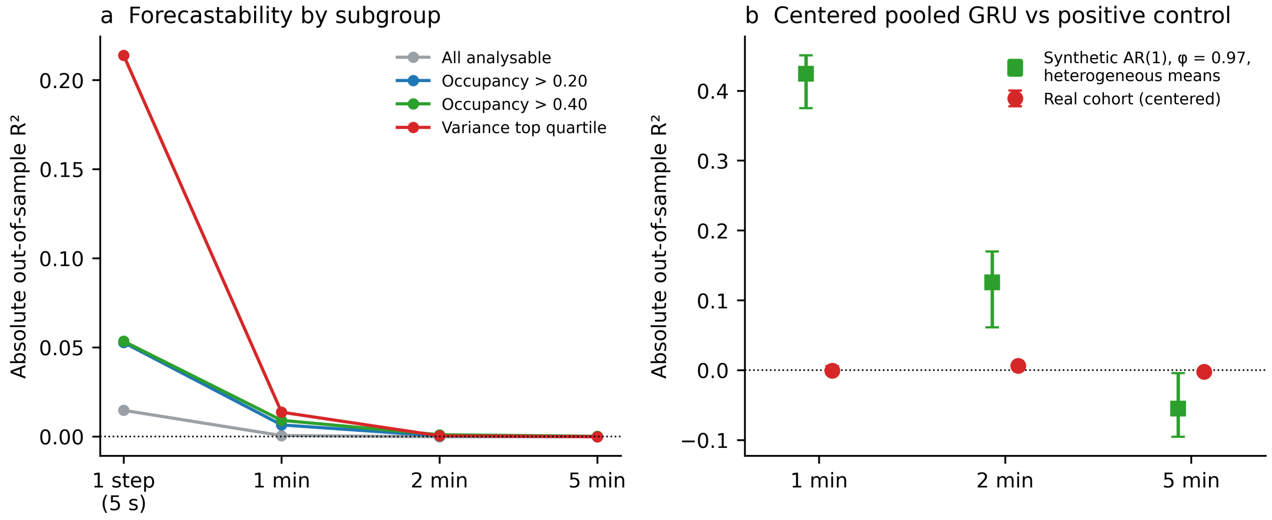


Figure S4. Robustness of the forecastability null (added in revision). (a) Absolute out-of-sample R squared by forecast horizon within mover subgroups; short-timescale (one-step) inertia rises with movement, reaching 0.21 in the top variance quartile, but multi-minute predictability collapses to zero in every subgroup. (b) Per-patient-centered pooled gated-recurrent-unit forecaster scored against each held-out patient's training-window mean: at or near zero at every horizon on the real cohort (red), but clearly positive at the 1- and 2-minute horizons on a synthetic heterogeneous-mean long-memory positive control processed by the identical pipeline (green; falling toward zero by 5 minutes as the finite window limits the recoverable signal), confirming the null is not a failure of model capacity. Centering each series on its own training-window mean removes the between-patient operating-level shift, so the panel tests temporal structure rather than that shift.

#### S4 Analysis Codebook

The analysis emits a single per-patient record with the fields below. Identifiers and intermediate arrays are retained only to allow exact reproduction of the predictability analysis without re-downloading raw signal.

- Operating level: per-window mean and median burst-suppression ratio.
- Non-saturated occupancy: fraction of grid cells inside each of the three bands.
- Regime geometry: burst and suppression dwell-time mean and coefficient of variation, regime-switch rate per hour.
- Predictability: absolute out-of-sample R squared and skill over persistence at the one-step, 1, 2, and 5 minute horizons; lag-1 autocorrelation.
- Within-window stability: augmented Dickey-Fuller probability, variance ratio, rolling-mean drift, and the binary stationary flag.
- Quality: clean-channel fraction, electromyographic-contamination index, number of grid cells, and the retained burst-suppression-ratio series.

#### S5 Code and Data Availability

The I-CARE v2.1 database is publicly available through PhysioNet under its data-use agreement. The analysis code, including the signal-retrieval and feature-extraction pipeline, the statistical analysis, and the figure generation, together with the deidentified derived per-patient feature table sufficient to reproduce every statistical figure and table without re-downloading raw EEG, is available at https://github.com/Alon-Gorenshtein/study_bsr_controllability under an open license (MIT for code, CC-BY for the derived table). No protected health information is contained in the derived feature tables, which hold only the quantitative measures described in S4.
